# Reduced trust in bodily sensations predicts suicidal ideation in hospitalized patients with major depression: an observational study

**DOI:** 10.1101/2024.12.18.24319219

**Authors:** Michael Eggart, Juan Valdés-Stauber, Bruno Müller-Oerlinghausen

## Abstract

**Background:** Major depressive disorder (MDD) is associated with maladaptive self-reported interoception, i.e., abnormal bodily self-experience. Although diminished body trusting predicts suicidal ideation, interoceptive measures have not been considered in depressed inpatients, whose suicide risk regularly peaks post-discharge. This study aims to explore interoceptive characteristics at admission that help identify inpatients at risk for suicidal ideation at discharge, thereby preventing fatal outcomes.

**Methods:** The observational study included 87 depressed inpatients providing self-ratings at both hospital admission (T0) and discharge (T1) on the following scales: Multidimensional Assessment of Interoceptive Awareness (MAIA-2); Beck Depression Inventory-II (BDI-II). A hierarchical logistic regression analysis estimated the longitudinal association between self-reported interoception (T0) and suicidal ideation (T1). The optimal cutpoints for predicting suicidal ideation were calculated using ROC curve analysis.

**Results:** Suicidal ideation was found in 17.24% patients at discharge, who reported lower baseline MAIA-2 *Trusting* scores than non-ideators (*p*=0.01). Diminished body trusting (*OR*=0.19), somatic comorbidity (*OR*=16.77), and baseline suicidal ideation (*OR*=24.01) significantly predicted suicidal ideation (T1). For body trusting, we estimated an optimal classification of subsequent suicidal ideation for the cutpoint≤2.33 (AUC=0.70 [95% CI 0.57, 0.83], sensitivity=0.87, specificity=0.44, positive predictive value=0.25, negative predictive value=0.94).

**Limitations:** Due to the exploratory nature of the study, the findings should be replicated in pre-registered trials with larger sample sizes.

**Conclusions:** Diminished body trusting is, with acceptable sensitivity, a significant predictor for post-treatment suicidal ideation in depressed inpatients. This finding emphasizes the importance of incorporating body-centered approaches into multimodal treatment strategies especially in inpatients under risk to prevent suicidal incidents.

## 1. Background

Major depressive disorder (MDD) is a common mental disorder associated with a substantial risk for suicide (Favril et al., 2023). Despite the rising numbers of antidepressant prescriptions, there is mounting evidence of a concurrent rise in suicide rates (Amendola et al., 2024). Antidepressants, particularly selective serotonin reuptake inhibitors (SSRIs), are suspected of inducing ego-dystonic suicidal thoughts, urges, and suicidal behaviors, especially shortly after initiation of treatment or following dose increases (Stübner et al., 2018). Psychiatric inpatient treatment is a high-risk period for suicide, as hospitalization often occurs when symptoms have reached a critical severity (Krause et al., 2020; Neuner et al., 2011). Research indicates that suicide risk peaks in the months following discharge (Chung et al., 2017; Forte et al., 2019; Goldacre et al., 1993; Ho, 2003; Park et al., 2013). However, reliable predictors for identifying hospitalized patients at risk are limited and differ between individuals only experiencing suicidal ideation and those with a history of suicide attempts (Lewitzka et al., 2017). Suicidal ideation – defined as “thoughts, ideas, or ruminations about the possibility of ending one’s life, ranging from thinking that one would be better off dead to formulation of elaborate plans” (WHO, 2024) – is a strong risk factor for suicidal mortality (Favril et al., 2023). Therefore, it is crucial to identify hospitalized patients at risk for persistent suicidal ideation early in their treatment. This calls for the exploration of new predictors of post-treatment suicidal ideation from the outset of hospitalization. In this context, abnormal body awareness may serve as a promising new predictor (Hielscher & Zopf, 2021).

Interoception encompasses the sensation, interpretation, and integration of signals “originating from within the body, providing a moment-by-moment mapping of the body’s internal landscape across conscious and unconscious levels” (p. 501) (Khalsa et al., 2018). These internal sensations, ranging from heartbeat and respiration to hunger and temperature, primarily serve to maintain homeostatic and allostatic regulation (Tsakiris & Critchley, 2016). The study of interoception has deep roots in phenomenology, physiology and neuroscience, and profound implications for mental health have recently gained significant attention (Cameron, 2001; Craig, 2002, 2009; Khalsa et al., 2018; Murphy et al., 2017; Tsakiris & Critchley, 2016). A state of dysfunctional interoception is increasingly considered as a fundamental component of MDD (Eggart, Lange, et al., 2019; Harshaw, 2015; Paulus & Stein, 2010): First, patient’s objective ability to accurately monitor their interoceptive states, known as *interoceptive accuracy* (Garfinkel et al., 2015), seems to be compromised in MDD. There is increasing evidence that affected subjects show diminished heartbeat perception accuracy (Eggart, Lange, et al., 2019) and reduced activation of the insular cortex during heartbeat perception tasks (Avery et al., 2014; Wiebking et al., 2010) – a state which potentially contributes to blunted positive affect intensity and decision-making difficulties (Furman et al., 2013). Second, the subjective attention to bodily sensations, known as *interoceptive sensibility* or *self-reported interoception* (Garfinkel & Critchley, 2013; Mehling, 2016), is abnormal in MDD: Core characteristics of MDD encompass altered pain perception (Thompson et al., 2016), increased somatic symptom burden (Kapfhammer, 2006), anxiety-driven attention styles directed towards unpleasant bodily cues (Flasinski et al., 2020; Zhou et al., 2024), emotion dysregulation linked with abnormal body awareness (Lyons, Strasser, et al., 2021; Zhou et al., 2022), and reduced confidence in bodily sensations (Dunne et al., 2021). From a phenomenological perspective, patient’s bodily feelings have been attributed to a ‘corporealization’ of the lived body (Fuchs, 2005; Fuchs & Schlimme, 2009), which is typified by sensations of constriction and oppression in the chest and abdomen, accompanied by feelings of alienation, heaviness, blockage, emptiness, paralysis, and passivity in the whole body (Lyons, Michaelsen, et al., 2021). These coenesthesias should not be solely construed as matters of well-being, since abnormal self-reported interoception has been recognized as a predictor for unfavorable treatment outcomes (Eggart et al., 2021; Eggart & Valdés-Stauber, 2021) and as a risk factor for residual symptoms of fatigue (Eggart et al., 2023a).

Recent studies have begun to shed light on the potential relationship between dysfunctional interoception and suicidality. Overall, a systematic review of the available evidence indicates that interoceptive impairments may be viable risk factors contributing to the emergence of suicidal ideation and behaviors (Hielscher & Zopf, 2021). The literature consistently demonstrates a robust association between diminished trust in bodily sensations and the occurrence of suicidal ideation (Duffy et al., 2018, 2020; Forkmann et al., 2019; Gioia et al., 2022; Rogers et al., 2018). Additionally, low confidence in the body is related to a history of suicide attempts (Duffy et al., 2018, 2020; Rogers et al., 2018). However, it is important to acknowledge the limitations present in most prior studies within this domain, particularly with regard to their use of cross-sectional research designs, and insufficient control for disease-specific confounding variables such as depression severity (Hielscher & Zopf, 2021). Furthermore, there is a notable lack of longitudinal studies that have specifically examined interoceptive predictors of suicidal ideation among inpatients diagnosed with MDD.

Therefore, we explored prospective associations between self-reported interoception in MDD inpatients at admission and the likelihood of reporting suicidal ideation upon psychiatric discharge, aiming to enable early identification of at-risk patients during the initial phase of hospital treatment. Furthermore, we aimed to delineate interoceptive baseline disparities in body trusting between inpatient groups exhibiting differentiated change patterns in suicidal ideation from admission to discharge.

## 2. Methods

During all stages of the research process, the declaration of Helsinki has been considered, and patients gave their written informed consent to participate in the study. The study was approved by the ethics committee of Ulm University (registration number: 13/17).

### 2.1 Procedure and Participants

This observational study included 87 participants who were consecutively admitted to a hospital ward specialized for the treatment of MDD in the Department of Psychiatry and Psychotherapy I of Ulm University (ZfP Südwürttemberg, Weißenau) meeting inclusion criteria (main diagnosis of major depression, ≥18 years, proficiency of the German language). Patients were excluded if they showed symptoms of psychosis, schizophrenia, intellectual disability, or substance abuse. Included participants were 47.57 (±10.64) years old, 49 (56.32%) patients were female, and 60 (68,97%) were diagnosed with recurrent depressive disorder (F33). Most patients (*n* = 79, 90.81%) fulfilled criteria for severe depression (F3x.2), 8 (9.20%) patients were diagnosed with moderate depression (F3x.1). Diagnoses were assessed by trained psychiatrists or clinical psychologists according to ICD-10 criteria (WHO, 1992).

The present study is part of a larger project investigating interoceptive predictors of treatment outcomes in inpatients suffering from MDD. A detailed description of study characteristics and treatment components has been published in a previous paper (Eggart & Valdés-Stauber, 2021) and will be briefly summarized here. The treatment followed a guideline-based approach and was not changed during the study period. All patients received psychotherapy (weekly sessions on the individual and group level), and most participants (*n* = 83, 95.40%) were treated with antidepressants. The antidepressant therapy was complemented by nursing interventions (e.g., crisis intervention, professional communication, relaxation techniques) and by exercise therapy. Study data were assessed within 48 hours after admission (T0)/discharge (T1) to/from hospital, respectively. The median treatment duration was 8 weeks (interquartile range: 6.50-10.00). Further details on participants’ characteristics are presented in Table 1.

**Table 1:**
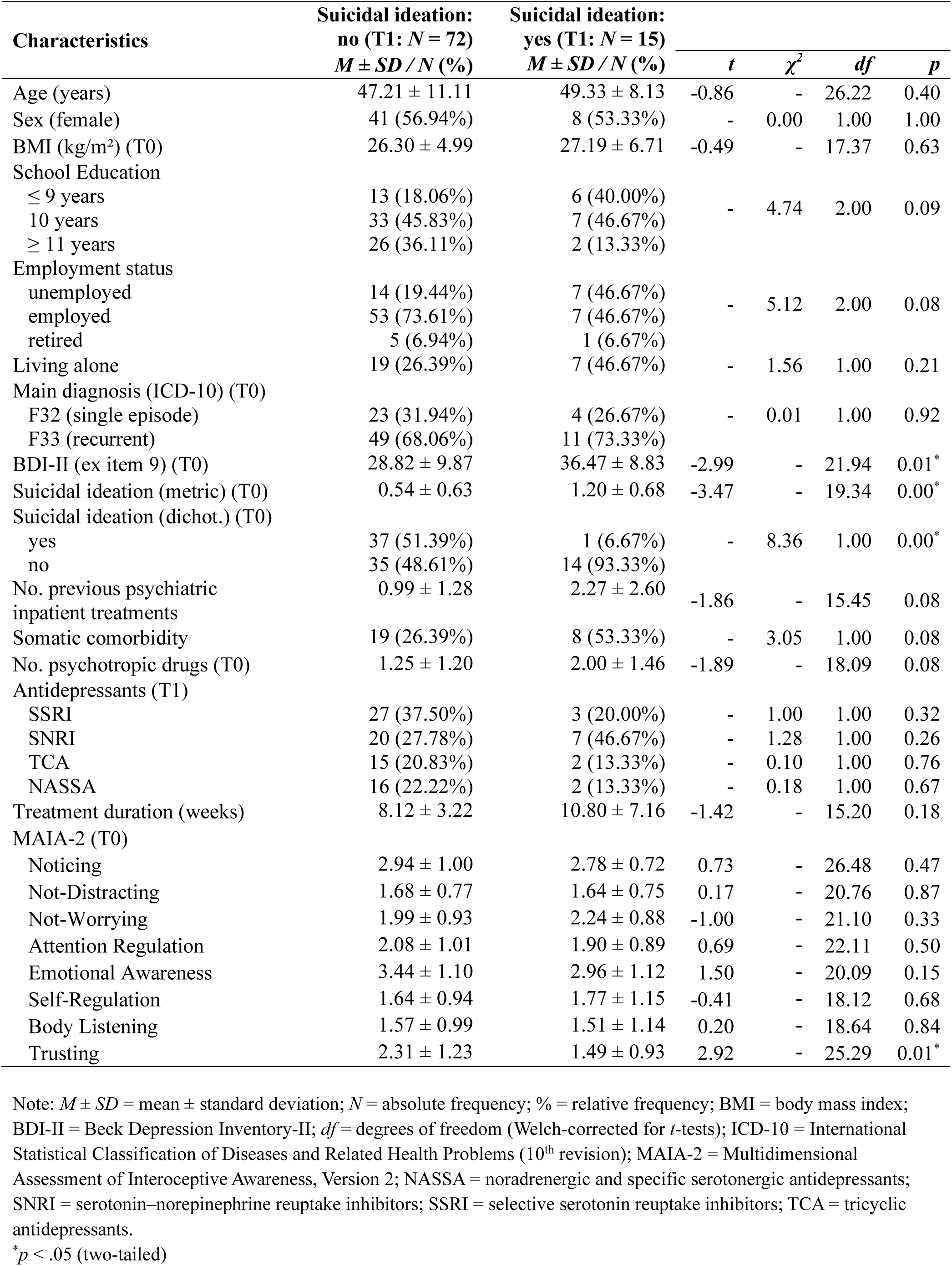
Participant characteristics (*N* = 87)

### 2.2 Measures

#### 2.2.1 Multidimensional Assessment of Interoceptive Awareness, Version 2 (MAIA-2)

The Multidimensional Assessment of Interoceptive Awareness, Version 2 (MAIA-2) is a self-rating scale which assesses differentiated aspects of self-reported interoception (Eggart et al., 2021; Mehling et al., 2018). The multidimensional questionnaire includes 37 items which are rated on a 6-point Likert scale (0 = “never”; 5 = “always”). The scales are averaged based on their respective items and are briefly described in the following (a sample item for each dimension and internal consistency reliability estimates for this study will be reported in brackets): *Noticing* (McDonald’s *ω* = 0.58; “I notice when I am uncomfortable in my body.”); *Not-Distracting* (*ω* = 0.67; “When I feel unpleasant body sensations, I occupy myself with something else so I don’t have to feel them.”); *Not-Worrying* (*ω* = 0.67; “When I feel physical pain, I become upset.”); *Attention Regulation* (*ω* = 0.87; “I can refocus my attention from thinking to sensing my body.”); *Emotional Awareness* (*ω* = 0.86; “I notice that my breathing becomes free and easy when I feel comfortable.”); *Self-Regulation* (*ω* = 0.76; “When I am caught up in thoughts, I can calm my mind by focusing on my body/breathing.”); *Body Listening* (*ω* = 0.77; “I listen for information from my body about my emotional state.”); *Trusting* (*ω* = 0.88; “I feel my body is a safe place.”) (Eggart et al., 2021). In previous research, the MAIA-2 demonstrated adequate reliability and construct validity (Mehling et al., 2012, 2018). The questionnaire also exhibited criterion validity in distinguishing between treatment response groups in depressed inpatients (Eggart et al., 2021). The strength of MAIA-2 is its ability to differentiate between clinically beneficial and maladaptive interoceptive states by assessing attention styles towards the body that are anxiety-driven or linked to self-regulatory benefits (Mehling, 2016). The questionnaire is frequently used in clinical samples and in the general population (Todd et al., 2020).

#### 2.2.2 Beck Depression Inventory-II (BDI-II)

The Beck Depression Inventory-II (BDI-II) is a frequently used self-rating scale which assesses severity of depression in patients diagnosed with MDD. The questionnaire is unidimensional and includes 21 items that are rated on a 4-point Likert scale (Hautzinger et al., 2006). A sample item of the BDI-II is (Beck et al., 1996): “Loss of Pleasure” (0 = “I get as much pleasure as I ever did from the things I enjoy”; 1 = I don’t enjoy things as much as I used to”; 2 = “I get very little pleasure from the things I used to enjoy”; 3 = “I can’t get any pleasure from the things I used to enjoy”). The items are summed up to assess overall depression severity. In the present study, we excluded item 9 from the sum score because this item was investigated as the outcome variable (please, see below). The internal consistency reliability of the BDI-II (ex item 9) in the present study was good (*ω* = 0.89). In previous research, the measure demonstrated adequate reliability and validity (Beck et al., 1996; Hautzinger et al., 2006).

#### 2.2.3 Suicidal ideation

Suicidal ideation was investigated by extracting item 9 from the BDI-II. The exact wording of this item is as follows (Beck et al., 1996): “Suicidal Thoughts or Wishes” (0 = “I don’t have any thoughts of killing myself.”; 1 = “I have thoughts of killing myself, but I would not carry them out.”; 2 = “I would like to kill myself.”; 3 = “I would kill myself if I had the chance.”). We dichotomized patients’ responses to construct a binary variable which classified patients who reported suicidal thoughts of any severity (0 = “no suicidal ideation”; 1-3 = “suicidal ideation”).

### 2.3 Data analysis

The statistical analysis was performed in R version 4.3.0 (R Core Team, 2023) including the following R packages: cutpointr 1.1.1, fmsb 0.7.5, ggpubr 0.6.0, margins 0.3.26.1, MBESS 4.8.1, OptimalCutpoints 1.1-5, performance 0.8.0, psych 2.1.9, and tidyverse 1.3.1. For all analyses, the significance level was a priori set to 5%. Differences between patient groups with and without suicidal ideation were estimated by using *t*-tests (metric variables) or Chi^2^-tests (categorial variables). In the analysis aimed at delineating interoceptive baseline disparities between inpatient groups with differentiated change patterns in suicidal ideation from admission to discharge, we included the MAIA-2 *Trusting* scale due to its significance in previous research in suicidology (Hielscher & Zopf, 2021) and conducted a one-way ANOVA followed by Tukey’s post-hoc tests for multiple comparisons. A hierarchical logistic regression analysis was run to estimate the association between self-reported interoception (T0) and suicidal ideation (T1). In the first block, the following control variables were included to account for potential confounding effects with self-reported interoception based on previous research (Eggart et al., 2023a; Eggart & Valdés-Stauber, 2021): age, sex, body mass index, somatic comorbidity. The regression model was also adjusted for depression severity (BDI-II ex item 9, T0) and suicidal ideation (T0). In a separate sensitivity analysis, we exclusively included the MAIA-2 *Trusting* scale in the second block to determine its single contribution to the model. The odds ratio (*OR*) and average marginal effects (*AME*) were reported as effect measures along with 95% confidence intervals (95% CI).

We conducted a receiver operating characteristic (ROC) curve analysis to identify the diagnostic cutpoint at admission which predicts suicidal ideation at discharge. In the ROC curve analysis, a cutpoint for the MAIA-2 *Trusting* scale (T0) was derived on the basis of Youden’s index by maximizing the sum of sensitivity and specificity (Youden, 1950). This cutpoint classified patients who reported suicidal ideation (= positive test) of any severity versus no suicidal ideation (= negative test) at the end of treatment. The area under the curve (AUC) was estimated by plotting the sensitivity (*y*-axis) against 1 –specificity (*x*-axis). The quality of classification, i.e. accuracy, was further investigated by estimating the proportion of correctly classified cases. We also checked the assumptions of ROC curve analysis requiring a minimal correlation of |*r*| ≥ .30 for the associated metric variables (King, 2011).

## 3. Results

Participants’ characteristics grouped by suicidal ideation status (T1) are shown in Table 1. The prevalence of patients (*n* = 87) with any suicidal ideation was 56.32% (*n* = 49) at baseline (T0) and 17.24% (*n* = 15) at discharge (T1). Patients with suicidal ideation at psychiatric discharge showed higher baseline severity of depression and higher burden of suicidal ideation as well as significantly lower confidence in the body.

Overall, suicidal ideation significantly decreased during the course of treatment (T0: 0.66 ± 0.68; T1: 0.18 ± 0.42), *t*(86) = 6.82, *p* < 0.01 (two-tailed). In detail, suicidal ideation of any severity developed in 1 (1.15%) patient (‘no_T0_→yes_T1_’), disappeared in 35 (40.23%) patients (‘yes_T0_→no_T1_’), remained in 14 (16.09%) patients (‘yes_T0_→yes_T1_’), and was not reported at any time point by 37 (42.53%) patients (‘no_T0_→no_T1_’). We explored whether baseline scores on the MAIA-2 *Trusting* scale differed between these groups after excluding the single patient who developed suicidal ideation during hospital treatment (Figure 1). There was a significant difference in body confidence between the three groups, *F*(2, 83) = 7.01, *p* < 0.01, 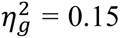 (large effect). The assumption of homoscedasticity was met by visually checking the Q-Q plot (not reported). In a post-hoc multiple comparison analysis, the Tukey HSD test revealed that baseline *Trusting* scores in patients who stopped with suicidal ideation during treatment (‘yes_T0_→no_T1_’: 1.91±1.08) were lower than in patients without any suicidal ideation at both time points (‘no_T0_→no_T1_’: 2.68±1.26), *ΔM* = -0.77 [95% CI -1.41, -0.13], *p_adj_* = 0.01. Accordingly, patients in whom suicidal ideation remained unchanged during treatment reported lower baseline *Trusting* scores (‘yes_T0_→yes_T1_’: 1.50±0.97) compared to the ‘no_T0_→no_T1_’ group, *ΔM* = -1.18 [95% CI -2.04, -0.33], *p_adj_* < 0.01. However, probably due to lack of statistical power, there was no significant difference between the ‘yes_T0_→yes_T1_’ and the ‘yes_T0_→no_T1_’ group, *ΔM* = -0.41 [95% CI -1.28, 0.45], *p_adj_* = 0.49.

**Figure 1.**
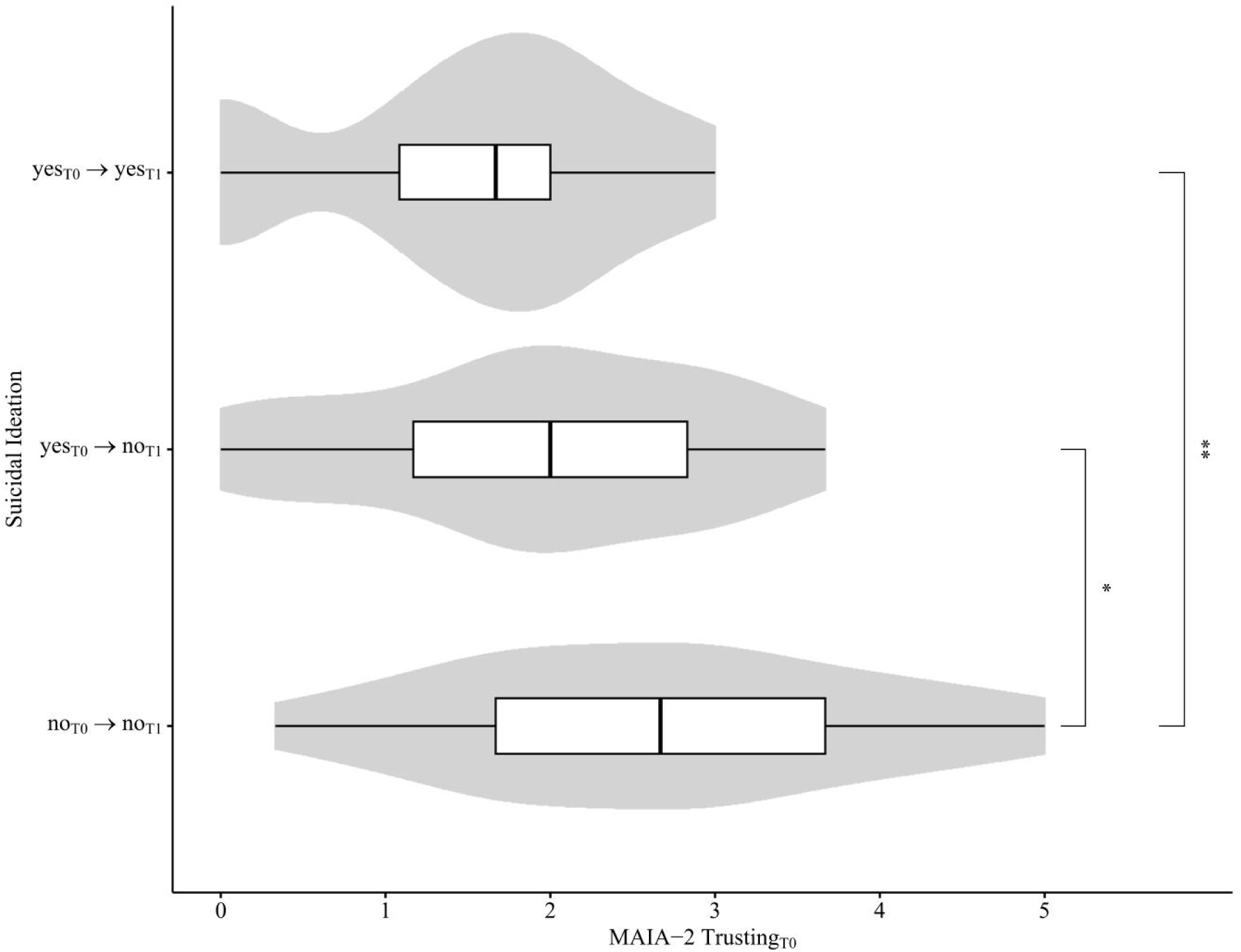
Difference in baseline body trusting between groups with differentiated change patterns in suicidal ideation from admission (T0) to discharge (T1). Patients who reported no suicidal ideation at any time point showed the highest baseline ratings in body trusting. The hybrid of a boxplot and a kernel density plot (grey) depicts the ratings on the MAIA-2 *Trusting* scale. The white boxplots show the median (vertical black line) and first/third quarter (lower/upper limit of the box) of the data. Asterisks indicate statistically significant differences (Tukey HSD test: \**p* < 0.05, \*\**p* < 0.01).

In a logistic regression analysis (Table 2), we identified low body trusting, *AME* = -0.13 [95% CI -0.21, -0.05], and the occurrence of somatic comorbidity, *AME* = 0.25 [95% CI 0.08, 0.41], as significant predictors of suicidal ideation over the time course, even after adjusting for baseline suicidal ideation that also showed a significant effect, *AME* = 0.20 [95% CI 0.08, 0.32]. Regarding self-reported interoception, this means that a one-unit increase on the MAIA-2 *Trusting* scale at baseline is associated with an averaged 13.01% decreased probability of experiencing suicidal ideation at the time of discharge. The statistical model has significantly improved after inclusion of the eight MAIA-2 scales, showing an increase of explained variance by 23.23% (Nagelkerke’s *R^2^*), χ^2^ (8) = 16.83, *p* = 0.03. Assumptions of logistic regression analysis were checked for the analyzed models, and multicollinearity was not identified (VIF < 10).

**Table 2:**
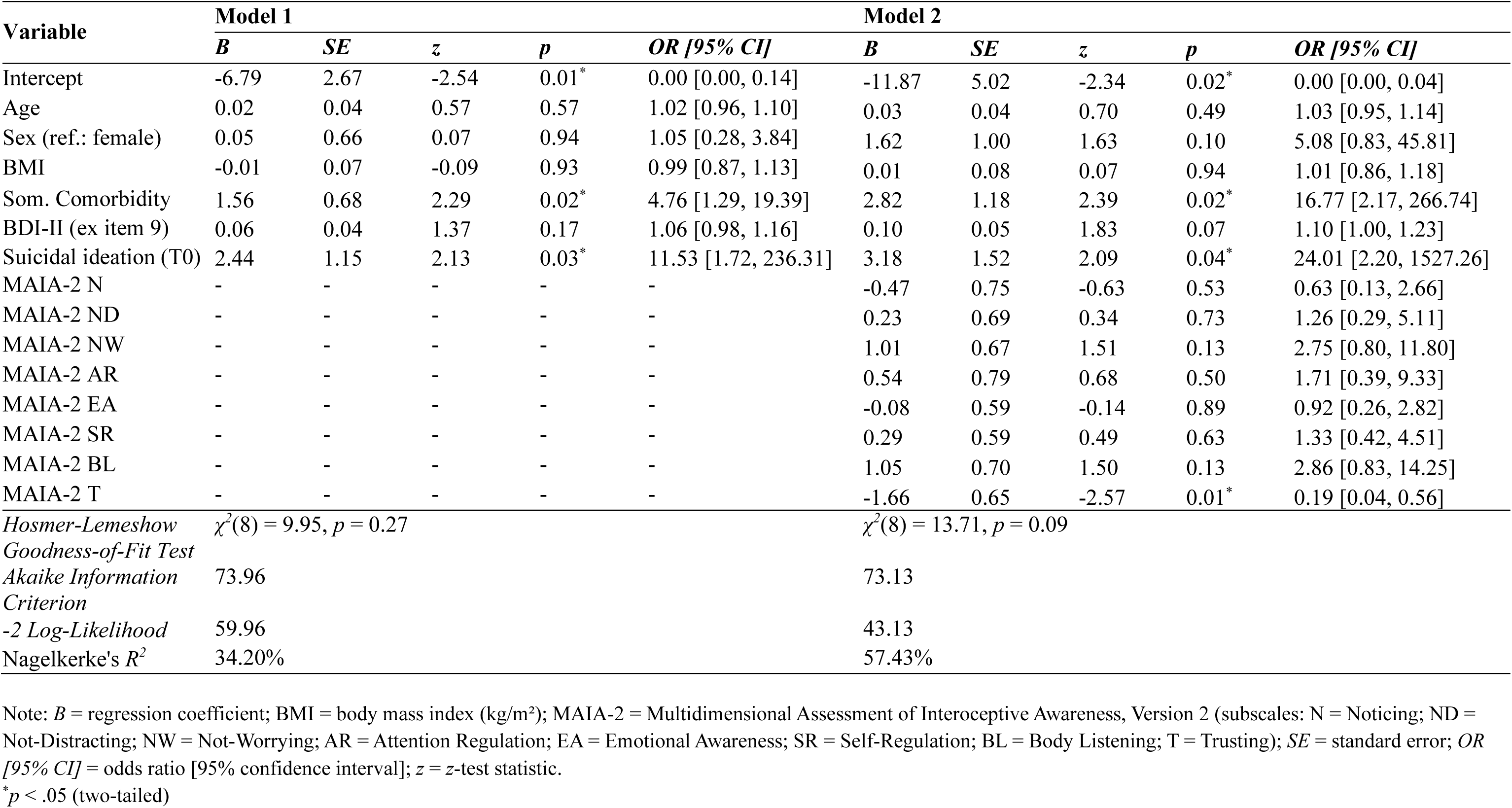
Prediction of suicidal ideation (T1) by baseline (T0) self-reported interoception (*N* = 87).

In an additional sensitivity analysis, we exclusively included the MAIA-2 *Trusting* scale in the hierarchical regression analysis to estimate its single contribution to the model and found a comparable significant effect, *B* = -0.76, *SE* = 0.38, *z* = -1.99, *p* < 0.05, *OR* = 0.47 [95% CI 0.20, 0.94], *AME* = -0.08 [95% CI -0.15, -0.01]. The explained variance increased by 6.82%, χ^2^ (1) = 4.61, *p* = 0.03.

The central assumption of ROC curve analysis was checked by showing a significant correlation between MAIA-2 *Trusting* (T0) and suicidal ideation (T1), *r* = -0.28 (95% CI - 0.46, -0.08), *p* < 0.01. Regarding the total sample, a ROC cutpoint of ≤2.33 on the *Trusting* scale (Figure 2) optimally classified patients under subsequent risk for suicidal ideation at discharge (AUC: 0.70 [95% CI 0.57, 0.83]; sensitivity: 0.87 [95% CI 0.60, 0.98]; specificity: 0.44 [95% CI 0.33, 0.57]; accuracy: 0.52). In this highly depressed sample, the positive predictive value, i.e. true positive/(true positive+false positive) = 13/(13+40), was 24.53% [95% CI 16.50, 74.79]. The negative predictive value, i.e. true negative/(true negative+false negative) = 32/(32+2), was 94.12% [95% CI 78.37, 96.31]. This means that, based on the prevalence of suicidal ideation (T1) = 17.24% in this sample, patients with a baseline *Trusting* score > 2.33 were at low risk for suicidal ideation, whereas every fourth patient with a score ≤ 2.33 showed suicidal ideation at discharge.

**Figure 2.**
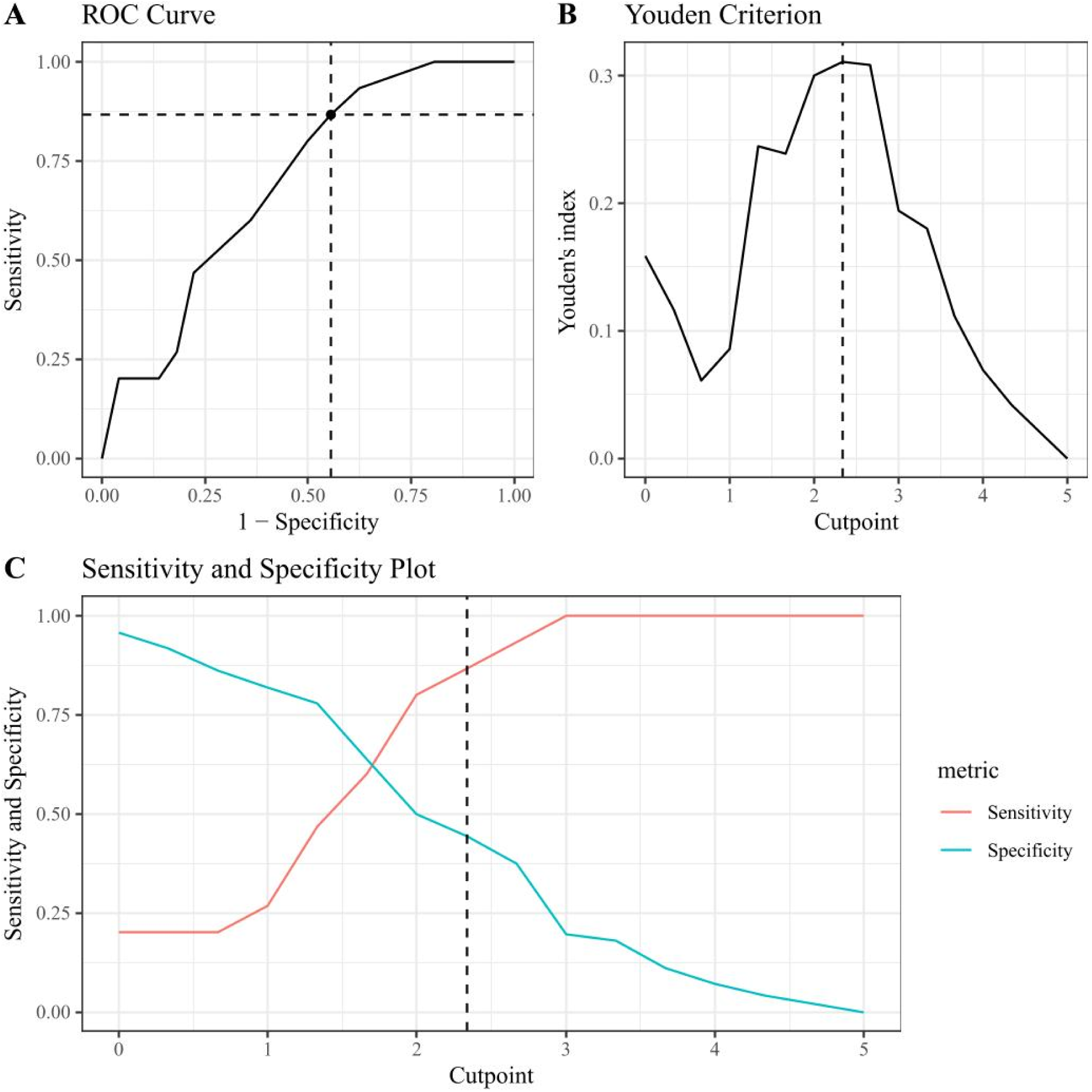
Results of ROC curve analysis establishing a binary classification of baseline MAIA-2 *Trusting* scores predicting suicidal ideation at discharge. **A.** The ROC curve shows the true-positive rate (sensitivity, *y*-axis) and the false-positive rate (1 – specificity, *x*-axis) for the optimal cutpoint (dashed line). **B.** The optimal cutpoint was extracted following the Youden criterion (Youden, 1950), Yl_c_ = max(Se_c_ + Sp_c_ − 1) = 0.31, which yielded the cutpoint *c* ≤ 2.33 (dashed line). There was no evidence of multiple cutpoints. **C.** Sensitivity and specificity plot for every possible baseline *Trusting* cut-point as a predictor of suicidal ideation. The dashed line shows the optimal cutpoint according to the Youden criterion.

## 4. Discussion

In the present study, we sought to identify interoceptive predictors of suicidal ideation in patients suffering from MDD. In summary, diminished body trusting, i.e. experiencing the body as unsafe and untrustworthy, after admission to hospital significantly predicted suicidal ideation at psychiatric discharge. Main findings and clinical implications for suicide prevention are discussed in the following sections.

Research on the prediction of suicidal ideation has so far yielded few conclusive results, particularly concerning prospective predictions over the time course of hospital treatment (Beck et al., 1989; Hawton et al., 2013). In this study, we identified, for the first time, a cutpoint on the *Trusting* subscale of the MAIA-2 that served as a significant predictor of subsequent suicidal ideation. The cutpoint demonstrated strong sensitivity, indicating that a substantial proportion of patients who reported suicidal ideation by the time of hospital discharge had a high probability of exhibiting subthreshold scores on the *Trusting* scale at baseline. However, the specificity of this cutpoint was suboptimal, leading to a higher rate of false-positive classifications by this diagnostic classifier. At first glance, the limitations of low specificity may seem disappointing, as it results in a low positive predictive value. However, it is important to consider that, given the current scarcity of predictors for suicidal ideation, even a predictor with limited classification accuracy is better than having no reliable predictors at all, which is currently the case. The cutpoint may help to identify patients at risk for suicidal ideation, enabling clinicians to tailor treatments for this high-risk group and better prepare for potential suicidal incidents in the critical period after hospital discharge – a time where suicidal risk is highest across all treatment stages (Chung et al., 2017; Forte et al., 2019; Goldacre et al., 1993; Ho, 2003; Krause et al., 2020; Park et al., 2013). However, research indicates that suicidal ideation and progression from suicidal thoughts to suicidal attempt is a distinct phenomenon with different predictors and mechanisms. While factors such as depression, hopelessness, and impulsivity are significant predictors of suicidal ideation, these variables do not consistently differentiate between individuals with suicidal ideation and suicide attempters (Klonsky et al., 2016). In summary, our results highlight the potential of interoceptive measures in predicting suicidal ideation, while emphasizing the need for improvements in the measure’s specificity by considering further factors to reduce false-positive rates.

Our findings align with recent reports by Gioia et al. (2022), who examined interoceptive predictors of suicidal ideation in a sample of participants without a clinical diagnosis of MDD using a longitudinal follow-up design. Their study revealed that diminished body trusting uniquely predicted both the presence and severity of suicidal ideation over a period of six months. However, unlike our study, the authors did not adjust for baseline depression severity. Furthermore, accumulating evidence from cross-sectional studies demonstrates similar associations between appraising bodily sensations as unsafe or untrustworthy and the presence of suicidal thoughts, thereby further supporting our findings (Duffy et al., 2020; Hielscher & Zopf, 2021; Perry et al., 2021; Rogers et al., 2018). Reduced body trusting has been suggested as an indicator of detachment from the body, a state that may contribute to an intensified desire for death (Belanger et al., 2023). Therefore, Smith et al. (2021) sought to enhance interoceptive dimensions, particularly body trusting, through an online intervention incorporating progressive muscle relaxation techniques, which effectively reduced outcomes related to suicidal ideation. These preliminary findings indicate that dysfunctional self-reported interoception may function not only as a risk factor for suicidal ideation but also as a promising target for interventions aimed at reducing the risk of suicide attempts or completions. While causal dynamics remain challenging to establish, psychological frameworks such as the concept of the “body-self” and the psychopathological construct of “depersonalization” warrant further exploration. Reduced body trusting could be clinically reinterpreted as depersonalization, i.e. a tendency not to take the bodily experience for granted but to focus on it perceptually, thereby awakening a gradual feeling of disconnection. Moreover, insights from philosophical and medical anthropology propose that experiential phenomena can be organized along fundamental pre-reflective dimensions, including “corporality” (Fuchs, 2018, 2021; Merleau-Ponty, 1945; Schmitz, 2011; Valdés-Stauber, 2024). In the context of certain physical and mental disorders, disruptions in this dimension may contribute to distorted self-perception and exacerbate psychopathological processes.

In hospitalized patients with MDD, it has been demonstrated that improvements in body trusting during treatment-as-usual are associated with a better response to therapy. This effect was particularly pronounced in women (Eggart & Valdés-Stauber, 2021). Furthermore, an experimental treatment involving ketamine in healthy individuals has demonstrated adverse effects on various aspects of self-reported interoception, by inducing a state of disembodiment and particularly by reducing body trusting (Kaldewaij et al., 2024). Consequently, study participants expressed a desire for any kind of touch or physical contact from significant others. These preliminary findings may provide new insights into the mechanisms that promote positive body awareness, which could potentially be enhanced through increased skin-to-skin contact (Kaldewaij et al., 2024). Clinical studies have also shown that massage therapy utilizing affective touch (i.e., gentle, slow, and rhythmic touch resembling effleurage techniques) can yield antidepressant, anxiolytic, analgesic, and hopelessness-reducing effects in individuals with MDD (Arnold et al., 2020; Baumgart et al., 2011, 2020; Hou et al., 2010; McGlone et al., 2024; Moyer et al., 2004; Müller-Oerlinghausen et al., 2004), probably through an interoceptive mechanism of action (Bohlen et al., 2021; Eggart, Queri, et al., 2019). Given this context, it is plausible that fostering positive tactile experiences could serve as a protective factor against suicidal thoughts or suicide, promoting body awareness, emotional regulation, a sense of safety, and social belongingness (Belanger et al., 2023; Heatley Tejada et al., 2020; Noone & McKenna-Plumley, 2022; Orbach, 2003; Orbach et al., 2006; Silvestri et al., 2023). This hypothesis warrants further investigation and builds upon key assumptions from social theories of suicide (Durkheim, 1897; Joiner, 2005), highlighting social isolation as a major risk factor (Motillon-Toudic et al., 2022). A sociological perspective suggests that interpersonal contact holds affective significance, corresponding to the anthropological dimension of “intercorporeality”. At a certain proximity, interpersonally relevant interactions may overlap with the dimension of “corporeality,” emphasizing the connection between relational and embodied experiences (Fuchs, 2013; Tanaka, 2015).

This study has several limitations that should be addressed in future research. The observational nature of this secondary data analysis limits definite causal interpretation, as it precludes control over potentially relevant and unknown confounders which interact with the MAIA-2 *Trusting* scale. To strengthen the validity of these findings, future preregistered studies with sufficient statistical power are needed, using comprehensive designs and detailed, multi-item suicidal self-report measures. Larger sample sizes would also enhance control over type II errors, particularly for interoceptive predictors. Furthermore, our reliance on a psychiatric inpatient sample introduces a selection bias, restricting the generalizability of the findings to other populations. However, this homogeneous sample also represents a strength, as it allows to estimate validated cutpoints relevant to typical psychiatric hospital settings, enhancing risk assessment in this population. Due to the exploratory nature of this study, we were unable to identify the underlying etiology of reduced body trusting in MDD, an area that warrants further investigation (Eggart et al., 2023b). Additionally, the study was underpowered to explore potential interactions between somatic comorbidity and low body confidence, meaning that we cannot exclude the possibility that the observed association between body confidence and suicidal ideation may be specific to patients with somatic comorbidities. Despite the aforementioned limitations, this study has the advantage of generating hypotheses that open a new direction for research into the phenomenon of suicidal ideation.

## 5. Conclusion

In this study, the level of self-reported body trusting at the onset of hospital treatment emerged as a significant predictor of suicidal ideation at discharge. Routinely assessing body trusting in patients with depression could thus provide healthcare providers with valuable insights, facilitating the identification of individuals at elevated risk for suicidal ideation. If evidence of reduced body trusting is present, the administration of SSRIs should be approached with heightened caution as these substances can potentially induce suicidal thoughts and behaviors (Stübner et al., 2018). Investigating the mechanisms by which interoception influences mental health outcomes may reveal promising therapeutic pathways, particularly by integrating psychological *and* body-centered treatments into patient care to reduce suicide risk. Thus, body trusting could serve not only as an important prognostic factor but also as a potential target for interoceptive therapies.

## Data Availability

The datasets used and/or analyzed during the current study are available from the corresponding author on reasonable request.

## Declarations

### Ethics approval and consent to participate

The study was approved by the ethics committee of Ulm University (registration number: 13/17). The principles of the Declaration of Helsinki were followed. Patients gave their written informed consent.

### Conflict of interest

The authors declare no conflict of interest.

### Funding

Not applicable.

## Notes

### Competing Interest Statement

The authors have declared no competing interest.

### Funding Statement

This study did not receive any funding.

## References

Amendola, S., Plöderl, M., & Hengartner, M. P. (2024). Suicide Rates and Prescription of Antidepressants. Crisis. 10.1027/0227-5910/a000941

Arnold, M. M., Müller-Oerlinghausen, B., Hemrich, N., & Bönsch, D. (2020). Effects of Psychoactive Massage in Outpatients with Depressive Disorders: A Randomized Controlled Mixed-Methods Study. Brain Sciences, 10(10), 676. 10.3390/brainsci10100676

Avery, J. A., Drevets, W. C., Moseman, S. E., Bodurka, J., Barcalow, J. C., & Simmons, W. K. (2014). Major depressive disorder is associated with abnormal interoceptive activity and functional connectivity in the insula. Biological Psychiatry, 76(3), 258–266. 10.1016/j.biopsych.2013.11.027

Baumgart, S. B.-E., Baumbach-Kraft, A., & Lorenz, J. (2020). Effect of Psycho-Regulatory Massage Therapy on Pain and Depression in Women with Chronic and/or Somatoform Back Pain: A Randomized Controlled Trial. Brain Sciences, 10(10), 721. 10.3390/brainsci10100721

Baumgart, S. B.-E., Müller-Oerlinghausen, B., & Schendera, C. F. G. (2011). Efficacy of Massage Therapy on Depression and Anxious Disorders as well as on Depressiveness and Anxiety as Comorbidity – A Systematic Overview of Controlled Studies. *Physikalische Medizin*, *Rehabilitationsmedizin*, Kurortmedizin, 21(04), 167–182. 10.1055/s-0031-1279760

Beck, A. T., Brown, G., & Steer, R. A. (1989). Prediction of eventual suicide in psychiatric inpatients by clinical ratings of hopelessness. Journal of Consulting and Clinical Psychology, 57(2), 309–310.

Beck, A. T., Steer, R. A., & Brown, G. K. (1996). Manual for the Beck Depression Inventory-II. Psychological Corporation.

Belanger, A. N., Timpano, K. R., Eng, G. K., Bragdon, L. B., & Stern, E. R. (2023). Associations Between Suicidality and Interoception in Obsessive-Compulsive Disorder. Journal of Obsessive-Compulsive and Related Disorders, 39. 10.1016/j.jocrd.2023.100844

Bohlen, L., Shaw, R., Cerritelli, F., & Esteves, J. E. (2021). Osteopathy and Mental Health: An Embodied, Predictive, and Interoceptive Framework. Frontiers in Psychology, 12, 767005.

Cameron, O. G. (2001). Interoception: The inside story—A model for psychosomatic processes. Psychosomatic Medicine, 63(5), 697–710. 10.1097/00006842-200109000-00001

Chung, D. T., Ryan, C. J., Hadzi-Pavlovic, D., Singh, S. P., Stanton, C., & Large, M. M. (2017). Suicide Rates After Discharge From Psychiatric Facilities: A Systematic Review and Meta-analysis. JAMA Psychiatry, 74(7), 694–702. 10.1001/jamapsychiatry.2017.1044

Craig, A. D. (2002). How do you feel? Interoception: the sense of the physiological condition of the body. Nature Reviews Neuroscience, 3(8), 655–666. 10.1038/nrn894

Craig, A. D. (2009). How do you feel—Now? The anterior insula and human awareness. Nature Reviews Neuroscience, 10(1), 59–70. 10.1038/nrn2555

Duffy, M. E., Rogers, M. L., Gallyer, A. J., & Joiner, T. E. (2020). Body Trust and Agitation: Pathways to Suicidal Thoughts and Behaviors. Archives of Suicide Research, 24(sup2), 236–250. 10.1080/13811118.2019.1592039

Duffy, M. E., Rogers, M. L., & Joiner, T. E. (2018). Body trust as a moderator of the association between exercise dependence and suicidality. Comprehensive Psychiatry, 85, 30–35. 10.1016/j.comppsych.2018.06.005

Dunne, J., Flores, M., Gawande, R., & Schuman-Olivier, Z. (2021). Losing trust in body sensations: Interoceptive awareness and depression symptom severity among primary care patients. Journal of Affective Disorders, 282, 1210–1219. 10.1016/j.jad.2020.12.092

Durkheim, É. (1897). *Le suicide: Étude de sociologie*. Ancienne Librairie Germer Baillière.

Eggart, M., Lange, A., Binser, M. J., Queri, S., & Müller-Oerlinghausen, B. (2019). Major Depressive Disorder Is Associated with Impaired Interoceptive Accuracy: A Systematic Review. Brain Sciences, 9(6), 131. 10.3390/brainsci9060131

Eggart, M., Queri, S., & Müller-Oerlinghausen, B. (2019). Are the antidepressive effects of massage therapy mediated by restoration of impaired interoceptive functioning? A novel hypothetical mechanism. Medical Hypotheses, 128, 28–32. 10.1016/j.mehy.2019.05.004

Eggart, M., Todd, J., & Valdés-Stauber, J. (2021). Validation of the Multidimensional Assessment of Interoceptive Awareness (MAIA-2) questionnaire in hospitalized patients with major depressive disorder. PloS One, 16(6), e0253913. 10.1371/journal.pone.0253913

Eggart, M., & Valdés-Stauber, J. (2021). Can changes in multidimensional self-reported interoception be considered as outcome predictors in severely depressed patients? A moderation and mediation analysis. Journal of Psychosomatic Research, 141, 110331. 10.1016/j.jpsychores.2020.110331

Eggart, M., Valdés-Stauber, J., Müller-Oerlinghausen, B., & Heinze, M. (2023a). Dysfunctional self-reported interoception predicts residual symptom burden of fatigue in major depressive disorder: An observational study. BMC Psychiatry, 23(1), 667. 10.1186/s12888-023-05168-y

Eggart, M., Valdés-Stauber, J., Müller-Oerlinghausen, B., & Heinze, M. (2023b). Exploring Associations between C-Reactive Protein and Self-Reported Interoception in Major Depressive Disorder: A Bayesian Analysis. Brain Sciences, 13(2), 353. 10.3390/brainsci13020353

Favril, L., Yu, R., Geddes, J. R., & Fazel, S. (2023). Individual-level risk factors for suicide mortality in the general population: An umbrella review. The Lancet Public Health, 8(11), e868–e877. 10.1016/S2468-2667(23)00207-4

Flasinski, T., Dierolf, A. M., Rost, S., Lutz, A. P. C., Voderholzer, U., Koch, S., Bach, M., Asenstorfer, C., Münch, E. E., Mertens, V.-C., Vögele, C., & Schulz, A. (2020). Altered Interoceptive Awareness in High Habitual Symptom Reporters and Patients With Somatoform Disorders. Frontiers in Psychology, 11, 1859. 10.3389/fpsyg.2020.01859

Forkmann, T., Volz-Sidiropoulou, E., Helbing, T., Druke, B., Mainz, V., Rath, D., Gauggel, S., & Teismann, T. (2019). Sense it and use it: Interoceptive accuracy and sensibility in suicide ideators. BMC Psychiatry, 19(1), 334. 10.1186/s12888-019-2322-1

Forte, A., Buscajoni, A., Fiorillo, A., Pompili, M., & Baldessarini, R. J. (2019). Suicidal Risk Following Hospital Discharge: A Review. Harvard Review of Psychiatry, 27(4), 209–216. 10.1097/HRP.0000000000000222

Fuchs, T. (2005). Corporealized and disembodied minds: A phenomenological view of the body in melancholia and schizophrenia. *Philosophy*, *Psychiatry*, & Psychology, 12(2), 95–107.

Fuchs, T. (2013). Depression, intercorporeality, and interaffectivity. Journal of Consciousness Studies, 20(7–8), 219–238.

Fuchs, T. (2018). Ecology of the Brain: The Phenomenology and Biology of the Embodied Mind. Oxford University Press.

Fuchs, T. (2021). In Defence of the Human Being: Foundational Questions of an Embodied Anthropology. Oxford University Press.

Fuchs, T., & Schlimme, J. E. (2009). Embodiment and psychopathology: A phenomenological perspective. Current Opinion in Psychiatry, 22(6), 570–575. 10.1097/YCO.0b013e3283318e5c

Furman, D. J., Waugh, C. E., Bhattacharjee, K., Thompson, R. J., & Gotlib, I. H. (2013). Interoceptive awareness, positive affect, and decision making in major depressive disorder. Journal of Affective Disorders, 151(2), 780–785. 10.1016/j.jad.2013.06.044

Garfinkel, S. N., & Critchley, H. D. (2013). Interoception, emotion and brain: New insights link internal physiology to social behaviour. Commentary on: „Anterior insular cortex mediates bodily sensibility and social anxiety“ by Terasawa et al. (2012). Social Cognitive and Affective Neuroscience, 8(3), 231–234. 10.1093/scan/nss140

Garfinkel, S. N., Seth, A. K., Barrett, A. B., Suzuki, K., & Critchley, H. D. (2015). Knowing your own heart: Distinguishing interoceptive accuracy from interoceptive awareness. Biological Psychology, 104, 65–74. 10.1016/j.biopsycho.2014.11.004

Gioia, A. N., Forrest, L. N., & Smith, A. R. (2022). Diminished body trust uniquely predicts suicidal ideation and nonsuicidal self-injury among people with recent self-injurious thoughts and behaviors. Suicide and Life-Threatening Behavior, 52(6), 1205–1216. 10.1111/sltb.12915

Goldacre, M., Seagroatt, V., & Hawton, K. (1993). Suicide after discharge from psychiatric inpatient care. The Lancet, 342(8866), 283–286. 10.1016/0140-6736(93)91822-4

Harshaw, C. (2015). Interoceptive dysfunction: Toward an integrated framework for understanding somatic and affective disturbance in depression. Psychological Bulletin, 141(2), 311–363. 10.1037/a0038101

Hautzinger, M., Keller, F., & Kühner, C. (2006). Beck Depressions-Inventar (BDI-II). Revision. Harcourt Test Services.

Hawton, K., Casañas i Comabella, C., Haw, C., & Saunders, K. (2013). Risk factors for suicide in individuals with depression: A systematic review. Journal of Affective Disorders, 147(1–3), 17–28. 10.1016/j.jad.2013.01.004

Heatley Tejada, A., Dunbar, R. I. M., & Montero, M. (2020). Physical Contact and Loneliness: Being Touched Reduces Perceptions of Loneliness. Adaptive Human Behavior and Physiology, 6(3), 292–306. 10.1007/s40750-020-00138-0

Hielscher, E., & Zopf, R. (2021). Interoceptive Abnormalities and Suicidality: A Systematic Review. Behavior Therapy, 52(5), 1035–1054. 10.1016/j.beth.2021.02.012

Ho, T.-P. (2003). The suicide risk of discharged psychiatric patients. The Journal of Clinical Psychiatry, 64(6), 702–707. 10.4088/jcp.v64n0613

Hou, W.-H., Chiang, P.-T., Hsu, T.-Y., Chiu, S.-Y., & Yen, Y.-C. (2010). Treatment effects of massage therapy in depressed people: A meta-analysis. The Journal of Clinical Psychiatry, 71(7), 894–901. 10.4088/JCP.09r05009blu

Joiner, T. (2005). Why People Die by Suicide. Harvard University Press.

Kaldewaij, R., Salamone, P. C., Enmalm, A., Östman, L., Pietrzak, M., Karlsson, H., Löfberg, A., Gauffin, E., Samuelsson, M., Gustavson, S., Capusan, A. J., Olausson, H., Heilig, M., & Boehme, R. (2024). Ketamine reduces the neural distinction between self- and other-produced affective touch: A randomized double-blind placebo-controlled study. Neuropsychopharmacology, 49, 1767–1774. 10.1038/s41386-024-01906-2

Kapfhammer, H.-P. (2006). Somatic symptoms in depression. Dialogues in Clinical Neuroscience, 8(2), 227–239.

Khalsa, S. S., Adolphs, R., Cameron, O. G., Critchley, H. D., Davenport, P. W., Feinstein, J. S., Feusner, J. D., Garfinkel, S. N., Lane, R. D., Mehling, W. E., Meuret, A. E., Nemeroff, C. B., Oppenheimer, S., Petzschner, F. H., Pollatos, O., Rhudy, J. L., Schramm, L. P., Simmons, W. K., Stein, M. B., … Zucker, N. (2018). Interoception and Mental Health: A Roadmap. Biological Psychiatry: Cognitive Neuroscience and Neuroimaging, 3(6), 501–513. 10.1016/j.bpsc.2017.12.004

King, M. T. (2011). A point of minimal important difference (MID): A critique of terminology and methods. Expert Review of Pharmacoeconomics & Outcomes Research, 11(2), 171–184. 10.1586/erp.11.9

Klonsky, E. D., May, A. M., & Saffer, B. Y. (2016). Suicide, Suicide Attempts, and Suicidal Ideation. Annual Review of Clinical Psychology, 12, 307–330. 10.1146/annurev-clinpsy-021815-093204

Krause, T. J., Lederer, A., Sauer, M., Schneider, J., Sauer, C., Jabs, B., Etzersdorfer, E., Genz, A., Bauer, M., Richter, S., Rujescu, D., & Lewitzka, U. (2020). Suicide risk after psychiatric discharge: Study protocol of a naturalistic, long-term, prospective observational study. Pilot and Feasibility Studies, 6(1), 145. 10.1186/s40814-020-00685-z

Lewitzka, U., Spirling, S., Ritter, D., Smolka, M., Goodday, S., Bauer, M., Felber, W., & Bschor, T. (2017). Suicidal Ideation vs. Suicide Attempts: Clinical and Psychosocial Profile Differences Among Depressed Patients: A Study on Personality Traits, Psychopathological Variables, and Sociodemographic Factors in 228 Patients. The Journal of Nervous and Mental Disease, 205(5), 361–371. 10.1097/NMD.0000000000000667

Lyons, N., Michaelsen, M. M., Graser, J., Bundschuh-Müller, K., Esch, T., & Michalak, J. (2021). Bodily Experience in Depression: Using Focusing as a New Interview Technique. Psychopathology, 54(3), 150–158. 10.1159/000514128

Lyons, N., Strasser, A., Beitz, B., Teismann, T., Ostermann, T., Anderle, L., & Michalak, J. (2021). Bodily Maps of Emotion in Major Depressive Disorder. Cognitive Therapy and Research, 45, 508–516. 10.1007/s10608-020-10195-0

McGlone, F., Uvnäs Moberg, K., Norholt, H., Eggart, M., & Müller-Oerlinghausen, B. (2024). Touch medicine: Bridging the gap between recent insights from touch research and clinical medicine and its special significance for the treatment of affective disorders. Frontiers in Psychiatry, 15, 1390673. 10.3389/fpsyt.2024.1390673

Mehling, W. E. (2016). Differentiating attention styles and regulatory aspects of self-reported interoceptive sensibility. Philosophical Transactions of the Royal Society of London. Series B, Biological Sciences, 371(1708). 10.1098/rstb.2016.0013

Mehling, W. E., Acree, M., Stewart, A., Silas, J., & Jones, A. (2018). The Multidimensional Assessment of Interoceptive Awareness, Version 2 (MAIA-2). *PLOS ONE*, *13*(12), 0208034. 10.1371/journal.pone.0208034

Mehling, W. E., Price, C., Daubenmier, J. J., Acree, M., Bartmess, E., & Stewart, A. (2012). The Multidimensional Assessment of Interoceptive Awareness (MAIA). PLOS ONE, 7(11), 48230. 10.1371/journal.pone.0048230

Merleau-Ponty, M. (1945). Phénoménologie de la perception. Gallimard.

Motillon-Toudic, C., Walter, M., Séguin, M., Carrier, J.-D., Berrouiguet, S., & Lemey, C. (2022). Social isolation and suicide risk: Literature review and perspectives. European Psychiatry, 65(1), e65. 10.1192/j.eurpsy.2022.2320

Moyer, C. A., Rounds, J., & Hannum, J. W. (2004). A Meta-Analysis of Massage Therapy Research. Psychological Bulletin, 130(1), 3–18. 10.1037/0033-2909.130.1.3

Müller-Oerlinghausen, B., Berg, C., Scherer, P., Mackert, A., Moestl, H.-P., & Wolf, J. (2004). Effects of slow-stroke massage as complementary treatment of depressed hospitalized patients. Results of a controlled study (SeSeTra). Deutsche Medizinische Wochenschrift, 129(24), 1363–1368. 10.1055/s-2004-826874

Murphy, J., Brewer, R., Catmur, C., & Bird, G. (2017). Interoception and psychopathology: A developmental neuroscience perspective. Developmental Cognitive Neuroscience, 23, 45–56. 10.1016/j.dcn.2016.12.006

Neuner, T., Hübner-Liebermann, B., Haen, E., Hausner, H., Felber, W., Wittmann, M., & Agate, F. (2011). Completed Suicides in 47 Psychiatric Hospitals in Germany – Results from the AGATE-Study. Pharmacopsychiatry, 44(7), 324–330. 10.1055/s-0031-1284428

Noone, C., & McKenna-Plumley, P. E. (2022). Lonely for Touch? A Narrative Review on the Role of Touch in Loneliness. Behaviour Change, 39(3), 157–167. 10.1017/bec.2022.12

Orbach, I. (2003). Suicide and the Suicidal Body. Suicide and Life-Threatening Behavior, 33(1), 1–8. 10.1521/suli.33.1.1.22786

Orbach, I., Gilboa-Schechtman, E., Sheffer, A., Meged, S., Har-Even, D., & Stein, D. (2006). Negative bodily self in suicide attempters. Suicide and Life-Threatening Behavior, 36(2), 136–153. 10.1521/suli.2006.36.2.136

Park, S., Choi, J. W., Kyoung Yi, K., & Hong, J. P. (2013). Suicide mortality and risk factors in the 12 months after discharge from psychiatric inpatient care in Korea: 1989–2006. Psychiatry Research, 208(2), 145–150. 10.1016/j.psychres.2012.09.039

Paulus, M. P., & Stein, M. B. (2010). Interoception in anxiety and depression. Brain Structure & Function, 214(5–6), 451–463. 10.1007/s00429-010-0258-9

Perry, T. R., Wierenga, C. E., Kaye, W. H., & Brown, T. A. (2021). Interoceptive Awareness and Suicidal Ideation in a Clinical Eating Disorder Sample: The Role of Body Trust. Behavior Therapy, 52(5), 1105–1113. 10.1016/j.beth.2020.12.001

R Core Team. (2023). *R: A language and environment for statistical computing. R Foundation for Statistical Computing*, *Vienna*, *Austria*. https://link.springer.com/content/pdf/10.1007/s00180-020-01034-7.pdf

Rogers, M. L., Hagan, C. R., & Joiner, T. E. (2018). Examination of interoception along the suicidality continuum. Journal of Clinical Psychology, 74(6), 1004–1016. 10.1002/jclp.22564

Schmitz, H. (2011). Der Leib. de Gruyter.

Silvestri, V., Giraud, M., Macchi Cassia, V., & Nava, E. (2023). Touch me or touch me not: Emotion regulation by affective touch in human adults. Emotion, 24(4), 913–922. 10.1037/emo0001320

Smith, A. R., Forrest, L. N., Perkins, N. M., Kinkel-Ram, S., Bernstein, M. J., & Witte, T. K. (2021). Reconnecting to Internal Sensation and Experiences: A Pilot Feasibility Study of an Online Intervention to Improve Interoception and Reduce Suicidal Ideation. Behavior Therapy, 52(5), 1145–1157. 10.1016/j.beth.2021.02.001

Stübner, S., Grohmann, R., Greil, W., Zhang, X., Müller-Oerlinghausen, B., Bleich, S., Ruther, E., Moller, H.-J., Engel, R., Falkai, P., Toto, S., Kasper, S., & Neyazi, A. (2018). Suicidal Ideation and Suicidal Behavior as Rare Adverse Events of Antidepressant Medication: Current Report from the AMSP Multicenter Drug Safety Surveillance Project. The International Journal of Neuropsychopharmacology, 21(9), 814–821. 10.1093/ijnp/pyy048

Tanaka, S. (2015). Intercorporeality as a theory of social cognition. Theory & Psychology, 25(4), 455–472. 10.1177/0959354315583035

Thompson, T., Correll, C. U., Gallop, K., Vancampfort, D., & Stubbs, B. (2016). Is Pain Perception Altered in People With Depression? A Systematic Review and Meta-Analysis of Experimental Pain Research. The Journal of Pain, 17(12), 1257–1272. 10.1016/j.jpain.2016.08.007

Todd, J., Barron, D., Aspell, J. E., Toh, E. K. L., Zahari, H. S., Khatib, N. A. M., & Swami, V. (2020). Translation and validation of a Bahasa Malaysia (Malay) version of the Multidimensional Assessment of Interoceptive Awareness (MAIA). PLOS ONE, 15(4), 0231048. 10.1371/journal.pone.0231048

Tsakiris, M., & Critchley, H. (2016). Interoception beyond homeostasis: Affect, cognition and mental health. Philosophical Transactions of the Royal Society B: Biological Sciences, 371, 1708. 10.1098/rstb.2016.0002

Valdés-Stauber, J. (2024). Phänomenologie des Abschieds vom Leben: Eine onto-anthropologische Annäherung mithilfe des Konstrukts „Antizipatorische Daseinsverabschiedung*“* [University of Augsburg]. https://opus.bibliothek.uni-augsburg.de/opus4/frontdoor/index/index/docId/115016

WHO. (1992). Manual of the International Classification of Diseases and Related Health Problems (10th edition). World Health Organization.

WHO. (2024). International Classification of Diseases for Mortality and Morbidity Statistics, Eleventh Revision (ICD-11). World Health Organization.

Wiebking, C., Bauer, A., Greck, M., Duncan, N. W., Tempelmann, C., & Northoff, G. (2010). Abnormal body perception and neural activity in the insula in depression: An fMRI study of the depressed „material me“. The World Journal of Biological Psychiatry, 11(3), 538–549. 10.3109/15622970903563794

Youden, W. J. (1950). Index for rating diagnostic tests. Cancer, 3(1), 32–35. 10.1002/1097-0142(1950)3:1%3C32::AID-CNCR2820030106%3E3.0.CO;2-3

Zhou, H., Liu, J., Wu, Y., Huang, Z., Wang, W., Ma, Y., Zhu, H., Zhou, Z., Wang, J., & Jiang, C. (2024). Unveiling the interoception impairment in various major depressive disorder stages. CNS Neuroscience & Therapeutics, 30(8), e14923. 10.1111/cns.14923

Zhou, H., Zou, H., Dai, Z., Zhao, S., Hua, L., Xia, Y., Han, Y., Yan, R., Tang, H., Huang, Y., Du, Y., Wang, X., Yao, Z., & Lu, Q. (2022). Interoception Dysfunction Contributes to the Negative Emotional Bias in Major Depressive Disorder. Frontiers in Psychiatry, 13. 10.3389/fpsyt.2022.874859

